# Fecal bile acid profiles predict recurrence in patients with primary *Clostridioides difficile* infection

**DOI:** 10.1101/2022.06.08.22276161

**Authors:** Benjamin H. Mullish, Laura Martinez-Gili, Elena Chekmeneva, Gonçalo D. S. Correia, Matthew R. Lewis, Verena Horneffer-Van Der Sluis, Julie A. K. McDonald, Alexandros Pechlivanis, Julian R. F. Walters, Emma L McClure, Julian R. Marchesi, Jessica R. Allegretti

**Author notes:** Corresponding author **Postal address:** Dr Jessica R Allegretti, Division of Gastroenterology, Hepatology and Endoscopy, Brigham and Women’s Hospital, 850 Boylston Street, Suite 201, Chestnut Hill, MA 02467, USA, **Email:**. Joint first authors (Benjamin H. Mullish, Laura Martinez-Gili).

## Abstract

1.

**Background:** Factors that influence recurrence risk in primary *Clostridioides difficile* infection (CDI) are poorly understood, and tools to predict recurrence are lacking. Perturbations in microbial-derived bile acids (BAs) contribute to CDI pathogenesis and may be relevant to primary disease prognosis.

**Aims:** To define stool bile acid profiles and microbial bile-metabolising functionality in primary CDI patients, and explore signatures predicting recurrence.

**Methods:** Weekly stool samples were collected from primary CDI patients from the last day of anti-CDI therapy until recurrence, or through eight weeks post-completion otherwise. Ultra-high performance liquid chromatography-mass spectrometry (UHPLC-MS) was used to profile bile acids, and bacterial bile salt hydrolase (BSH) activity was measured to determine primary BA deconjugation capacity. Multivariate and univariate models were used to define differential BA trajectories in recurrers *versus* non-recurrers, and assess fecal bile acids as predictive markers for recurrence.

**Results:** Twenty (36%) out of 56 patients (median age 57, 64% male) recurred, with 80% of recurrence occurring within the first nine days post-antibiotic treatment. Principal component analysis (PCA) of stool bile acid profiles demonstrated clustering of samples by recurrence status and post-treatment time point. Longitudinal fecal bile acid trajectories in non-recurrers showed a recovery of secondary bile acids and their derivatives in non-recurring patients that was not observed in recurrers. BSH activity increased over time amongst patients who did not relapse (β= 0.056; likelihood ratio test *p*=0.018). A joint longitudinal-survival model identified five stool bile acids with AUROC > 0.73 for prediction of recurrence within nine days post-CDI treatment.

**Conclusions:** Gut bile acid metabolism dynamics differ in primary CDI patients between those who develop recurrence versus those who do not. Individual bile acids show promise in primary CDI patients as potential novel biomarkers to predict CDI recurrence.

## 2. Introduction

*Clostridioides difficile* infection (CDI) continues to present a considerable global disease burden, with an estimated annual incidence of 462,100 cases in the USA alone on latest assessment, and this trend is expected to increase as antimicrobial resistance is predicted to grow worldwide [1]. While a growing proportion of cases appear to be community-acquired [2], CDI remains the major cause of hospital-acquired gastrointestinal infection [3], leading to increased hospitalization time [4], clinical complications and mortality [5]. Furthermore, CDI is associated with considerable healthcare expenditure, equating to $1.5 billion annually within the USA [6].

Recurrent CDI remains a major clinical challenge. A key clinical dilemma in the management of CDI patients is prediction of the risk of recurrence caused by either re-exposure to *C. difficile* or reactivation of dormant spores within vulnerable patients. The rate of recurrence within eight weeks following treatment for a primary episode of CDI is 15-25%, and rises as high as 40-60% for patients experiencing further recurrences [7,8]. Updated CDI clinical guidelines recognise that the risk of recurrence in primary CDI patients may influence the preferred management approach and the need for preventative strategies, such as the potential benefit for bezlotoxumab in primary CDI patients with higher recurrence risk compared to those with lower risk [9,10]. However, at present, limited tools exist for the prediction of CDI recurrence [11], with risk of recurrence estimation based upon the use of clinical criteria (including age, immunocompromise, and severity of CDI at diagnosis [9,12]). The dynamic assessment of biomarkers could be important not only to stratify low- and high-risk patients, but also to further understand the mechanisms underlying recurrence.

One such biological area of interest relates to the contribution of gut microbiota-bile acid interactions to the pathogenesis of CDI [13]. Prior antibiotic exposure is well-established as the major risk factor for CDI [14] and entails a loss of microbial community members possessing bile-metabolising enzymes (including bile salt hydrolases (BSHs) and 7-α-dehydroxylase) [15–18]. More specifically, the antibiotic-exposed gut develops enrichment in primary bile acids (including taurocholic acid (TCA), a major pro-germinant trigger to *C. difficile* [19]), and loss of secondary bile acids, that have established roles in restricting the growth of *C. difficile* and its toxin activity [20,21] and in modulating regulatory T cell activity [22]. In addition, CDI is also characterised by reduced activity in the farnesoid X receptor (FXR)-fibroblast growth factor (FGF) axis, important for bile acid homeostasis [23]. Fecal bile acid profiles have shown promise for differentiating non-CDI diarrhea from CDI-related diarrhea [24].

Our recent analysis of clinical factors in a cohort of patients experiencing a first episode of uncomplicated CDI, demonstrated that primary diagnosis of CDI via toxin enzyme immunoassay (EIA) and treatment with metronidazole were both factors increasing the risk of recurrence [25]. Extending upon this work, we here present an analysis of longitudinal fecal bile acid dynamics after anti-CDI therapy cessation until recurrence or until 8 weeks post-therapy of patients within this cohort, with the joint aims of better delineating differences in microbiome-bile acid interactions in patients with primary CDI who recur or not, and in identifying potential biomarkers that may predict future CDI recurrence.

## 3. Methods

### 3.1. Patient cohort

This cohort has been previously-described [25]. In brief, all participating patients were recruited from the inpatient service at Brigham and Women’s Hospital (BWH; Boston, USA) as well as two surrounding community hospitals (Brigham and Women’s Faulkner Hospital and Newton Wellsley Hospital). Potentially eligible patients were identified by daily reports of patients with positive stool tests for *C. difficile* provided by the BWH Clinical Microbiology Laboratory. *C. difficile* infection was defined as the presence of diarrhea, positive laboratory tests (i.e. glutamate dehydrogenase (GDH) and EIA toxin or polymerase chain reaction for toxin B, depending upon the testing methods used at the associated hospital laboratory), and clinician decision to treat the patient for CDI. Primary CDI was defined as no prior episodes of CDI within the prior six months. Patients who were excluded were: patients with inflammatory bowel disease; patients with inherited or acquired immunodeficiencies; severe or fulminant CDI, as diagnosed by IDSA and/or ACG guidelines [9,10]; or the need for ongoing non-CDI antibiotic use that continued past the CDI antibiotic course.

Participating patients were recruited at the time of CDI diagnosis and samples collected through eight weeks post-completion of their anti-CDI therapy to assess for recurrence, with clinical assessments regarding presence or absence of diarrhea occurring at each time point. Active stool collection began one week after completing anti-CDI antibiotics; stool was collected up to twice a week for the first two weeks post-therapy completion, and then weekly through to week eight. Recurrence was suspected if participants developed diarrhea (Bristol stool scale 6-7) and at least three bowel movements daily for three days; if these criteria were met, stool was assayed for *C. difficile* via GDH/EIA, and patients were considered to have had a recurrence if both assays were positive. While 75 patients were part of the original clinical cohort [25], only 56 had provided post-treatment serial stool samples, and hence the present study has an effective *n*=56. The Institutional Review Board (IRB) of Brigham and Women’s Hospital gave ethical approval for this work. In addition, a UK National Research Ethics Center (13/LO/1867) also gave ethical approval for this.

### 3.2. Bile acid profiling

Fecal samples were analysed using ultra-high performance liquid chromatography-mass spectrometry (UHPLC-MS). The protocols used for fecal extract preparation [26] and spectra acquisition [18,27] were as previously-described.

Fecal extracts were prepared by extracting from lyophilized dried fecal samples using a 2:1:1 (vol) mixture of water, acetonitrile and 2-propanol to 10mg/mL, via use of a Biospec bead beater with 1.0⍰mm Zirconia beads followed by centrifugation (16,000×*g*, 20⍰min) and filtering the supernatant through 0.45⍰μm microcentrifuge filters (Costar, Corning). Pooled study reference (SR) samples, used as quality control of the profiling data and to monitor assay performance, were prepared using equal parts of the fecal filtrates. In addition, for assessment of linearity of analyte response [28], a series of SR sample dilutions was created by diluting with ice-cold LC-MS grade methanol to the concentrations of 100%, 80%, 60%, 40%, 20%, 10%, 1% and analyzed at the start and end of each set of sample analyses.

Study and pooled SR samples were prepared for the UHPLC-MS profiling by aliquoting 75 µL of filtered fecal extracts onto a 96-well plate and adding 75 µL of LCMS grade water and 75 µL of internal standard (IS) solution, followed by the addition of 75 µL of water to each well for study samples, and 75 µL of ice-cold LC-MS grade methanol to each well for study samples. Samples were mixed for two minutes on a plate mixer (1400 rpm at 2-8 °C), incubated at -20 °C for 4 hours, then mix briefly and centrifuged at 3486x*g* for 10 minutes at 4 °C. Aliquots of 125 µL of clear supernatants of each sample were carefully transferred to an analytical 96-well plate that was heat sealed and placed into the autosampler maintained at 4 °C for the analysis. Blank samples were prepared in the same way but with an empty tube (i.e. processing neat extraction solution through all steps, including the filter, allowing tracing back of contamination peaks if present).

ACQUITY UHPLC-MS (Waters Ltd., Elstree, UK) coupled to a Xevo G2-S Q-ToF mass spectrometer with an electrospray ionization source operating in negative ion mode (ESI–) (Waters, Manchester, UK) was used for bile acids profiling. LC separation was conducted on a ACQUITY BEH C8 column (1.7 μm, 100 mm × 2.1 mm) maintained at 60 °C. A gradient was applied consisting of 10:1 water:acetonitrile, 1 mM ammonium acetate, pH 4.15 (A) and 1:1 isopropanol: acetonitrile (B). Details of the linear gradient method 90% A to 65% are described elsewhere [18]. Injection volume was of 5 µL.

Mass spectrometry parameters were as follows: capillary voltage was set at 1.5 kV, cone voltage at 60 V, source temperature at 150 °C, desolvation temperature at 600 °C, desolvation gas flow at 1000 L/h, and cone gas flow at 150 L/h. Masslynx software (Waters, Manchester, U.K.) was used for data acquisition and visual inspection.

A total of 81 bile acid authentic chemical standards were used to help annotation of endogenous bile acids in fecal samples. The standards were split into eight mixtures prepared in 1:3 water: methanol mixture and analyzed in the beginning and the end of the run. The standards were also spiked into pooled SR sample analyzed across the run. This helped monitor any potential retention time shifts for each bile acid species and to determine retention time windows (regions of interest) used as an input together with *m/z* values for the targeted extraction and integration of annotated bile acids using peakPantheR package [27].

Waters .RAW LC-MS data files were converted to .mzML format using Proteowizard msconvert [29], with removal of signals with less than 100 ion counts. A total of 81 bile acid species were annotated using authentic reference standards to determine their retention times and their relative abundance integrated with the R package, peakPantheR [30]. To adjust for spectra intensity decay along the run, each feature was divided by a LOWESS curve fitted on the SR samples’ intensity [31]. Features with a coefficient of variation greater than 30% in normalised SR samples, with a Pearson correlation with dilution factor below 0.7 (estimated using a dilution series of pooled SR samples), or below LOD in more than 20% of study samples were discarded. Drift-correction and feature filtering were performed with the nPYc-Toolbox [32]. Features were log-transformed and zeros imputed using impute.QRILC from the *imputeLCMD* R package. For statistical analyses, features were also mean-centered.

### 3.3. Bile salt hydrolase activity assays

A spectrophotometry-based assay was used to assess the activity of microbial bile salt hydrolase (BSH) in a subset of stool samples from the cohort, as previously-described [26].

### 3.4. Data analysis and statistics

Statistical analyses were performed in R. Principal component analysis (PCA) of the processed UHPLC-MS bile acid profiling data was performed using the *ropls* R package[33]. Bile acid longitudinal trajectories were fitted using mixed effects models with the R package *nlme*, stratifying samples by recurrence, with time (transformed as square root of days for ease of modelling and visualisation; **Supplementary Figure 1**) as a co-variate and subject as random intercept and time slope effect. For each feature, log-likelihood ratio tests of nested models were used to determine the addition of non-linear (squared and cubic) time terms. Area under the curve (AUC) of subject-specific fitted bile acid trajectories up to the maximum timepoint of recurrence was calculated with *pracma* function trapz and compared across recurring and non-recurring patients using a Mann-Whitney test, with Benjamini-Hochberg false discovery rate correction (FDR) for multiple testing applied. FDR-adjusted P-values (*P*_*adj*_) < 0.1, corresponding to a 10 % false discovery rate, were considered significant.

To assess if bile acids contributed to recurrence, a Bayesian method was used to jointly model bile acid longitudinal changes with the hazard risk of recurrence using jointModelBayes function in the JMbayes package [34] with default parameters. The joint survival model included age, gender, previous antibiotics (yes/no), *C. difficile* antibiotic type (vancomycin/metronidazole) and diagnostic test (immunoassay/PCR) as covariates. Mean area under the receiver operating characteristic curve (AUROC) was calculated for each feature with the rocJM method from JMbayes package, using 1,000 Monte Carlo permutations of training sets randomly built with 80% of recurrers’ and 80% of non-recurrers’ samples and the remaining 20% of samples as test set. Three sets of ROC curves were built using only one, two or three first timepoints of longitudinal bile acid measurements to predict recurrence within nine days post-treatment.

## 4. Results

### 4.1. Participant details

Key clinical details of the 56 included patients are presented in **Table 1**. No patients had received treatment with ursodeoxycholic acid, and two patients were treated with bile acid sequestrants (one with colestipol, one with cholestyramine). Out of the 56 patients, 20 (36%) developed recurrence, with 80% of recurrence occurring within the first nine days post-antibiotic treatment.

**Table 1:** Clinical details of study participants.

| **Variable Name** | **n = 56** |
| --- | --- |
| Age (Mean ± SD) | 57.7 ± 16.3 |
| Male N,% | 36 (64.3%) |
| Race N,% |  |
| Black | 9 (16%) |
| White | 41 (73.2%) |
| BMI (Mean ± SD) | 28 ± 6.5 |
| Received Antibiotics prior to Diagnosis N,% | 40 (71.4%) |
| Prior PPI use N,% | 27 (48.2%) |
| History of Cirrhosis N,% | 3 (5.3%) |
| Dietary restrictions N,% |  |
| No | 52 (92.8%) |
| Vegan | 1 (1.7%) |
| Vegetarian | 2 (3.5%) |
| Gluten Free | 2 (3.5%) |
| Lactose Free | 1 (1.7%) |
| Smoking status |  |
| Never | 34 (60.7%) |
| Former | 20 (35.7%) |
| Current | 2 (3.6%) |
| Diagnosis of Irritable Bowel Syndrome N,% | 6 (10.7%) |
| Baseline diarrhea or constipation N,% |  |
| No | 42 (75%) |
| Diarrhea | 6 (10.7%) |
| Constipation | 6 (10.7%) |
| Both | 2 (3.6%) |
| Baseline Bristol Score (Mean ± SD) | 3.2 ± 1.3 |
| Ursodeoxycholic acid Use N,% | 0 (0.0%) |
| Cholestyramine Use N,% | 1 (1.7%) |
| Colestipol Use N,% | 1 (1.7%) |
| CDI Treatment Regimen |  |
| Metronidazole | 14 (25%) |
| Vancomycin | 42 (75%) |
| Other current antibiotics (not for *C. difficile*) | 0 (0%) |
| Test Used for Diagnosis N,% |  |
| PCR | 22 (39.2%) |
| EIA Toxin | 34 (60.7%) |
| **Baseline Lab Values (Mean ± SD)** |  |
| WBC | 9.8 ± 5.3 |
| Hct | 36.1 ± 5.6 |
| Plts | 237.6 ± 103.6 |
| ALT | 44.4 ± 91.6 |
| AST | 41.3 ± 89.2 |
| Alkaline Phosphatase | 81.9 ± 40.0 |
| T. Bilirubin | 0.6 ± 0.4 |
| BUN | 20.3 ± 23.3 |
| Cr | 1.9 ± 5.2 |
| PT | 22.8 ± 29.4 |
| INR | 1.3 ± 0.31 |

### 4.2. Timepoint since diagnosis and recurrence status influence fecal bile acid profile in primary CDI

In total, 71 bile acids were detected and relatively quantified in the peakPantheR dataset (see **Supplementary Table 1** for full list of bile acids).

We first sought to explore the overall impact of recurrence status and post-treatment sampling time upon fecal bile acid profiles; as such, sample profiles were first visualised using multivariate statistical analysis. On principal component analysis (PCA) of the annotated bile acid profiles of all included samples and time points (*n*=273), samples clustered mostly according to timepoint of sampling and by recurrence status of the patient (**Figure 1A**). Individual trajectories showed that patients who did not recur had more pronounced dynamic changes in their fecal bile acid composition than those who recurred (**Figure 1B**); this higher temporal variability in non-recurrers was reflected in the comparison of mean Euclidean distance of within-individual longitudinal samples collected the first nine days post-treatment, which showed a higher - but not significant - dissimilarity in bile acid composition across time in non-recurrers (Wilcoxon *p* = 0.13; **Figure 1C**).

**Figure 1:**
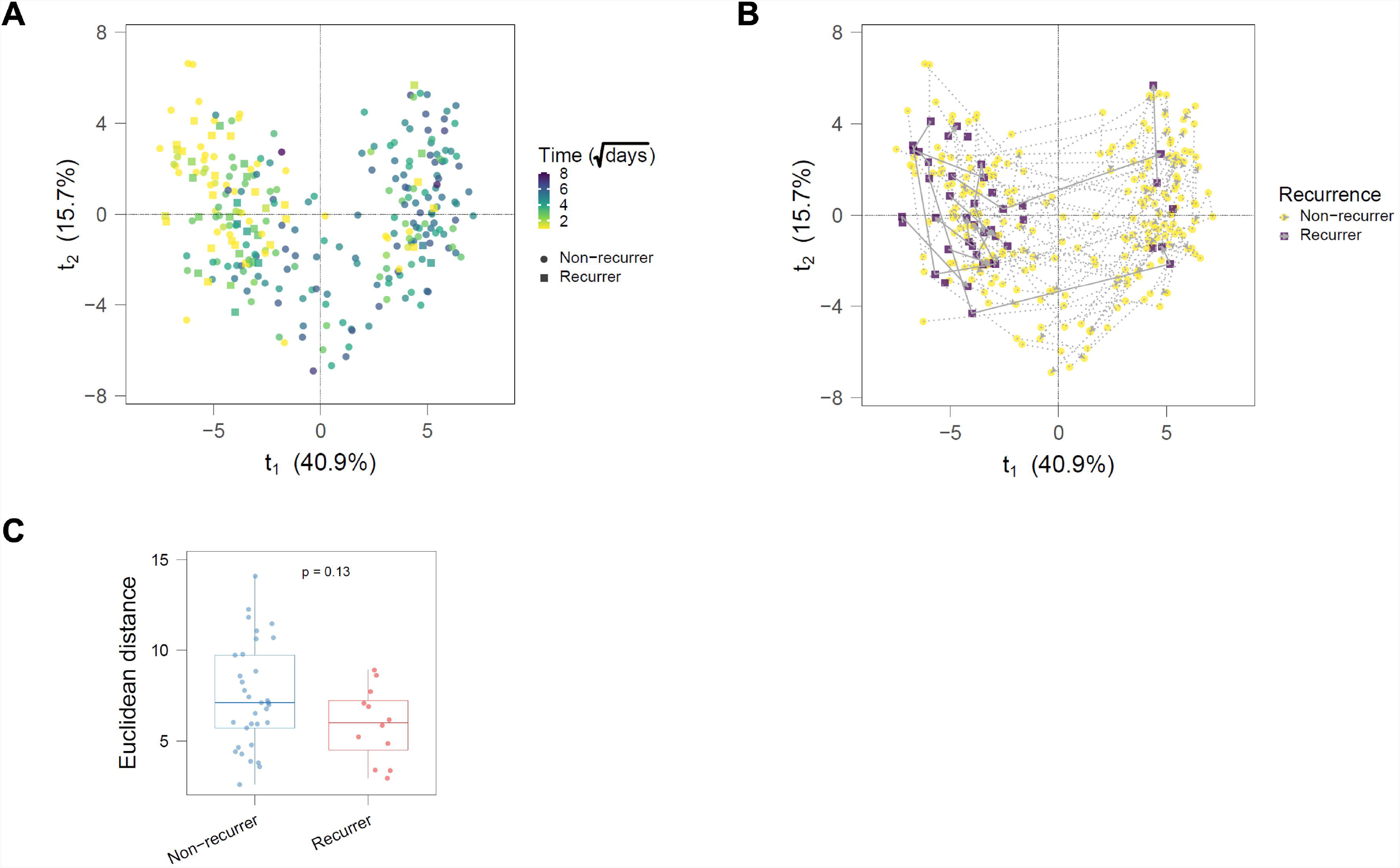
Initial multivariate statistical analysis of fecal bile acid profiles obtained from patients with primary CDI. Principal component analysis (PCA) of annotated bile acid relative intensities for all included samples (*n*=273), as coloured by time point of collection (A) or recurrence status (B). C) Average within-individual Euclidean distance of bile acid composition across timepoints for participants with more than one sample collected up to the first 9 days post-treatment (n= 45; 12 recurrers and 33 non-recurrers). *p* value was calculated using Wilcoxon test.

### 4.3. Recovery of microbial bile acid metabolism and restoration of the pre-morbid bile acid milieu occurs in primary CDI patients without recurrence

To explore the temporal changes in bile acid metabolism in patients recovering from primary CDI – and how recurrence may impact upon this – we compared the longitudinal trajectories (across days to weeks) of fecal bile acids for the 20 patients experiencing recurrence *versus* those 36 with no recurrence (**Figure 2A, B** and **C**).

**Figure 2:**
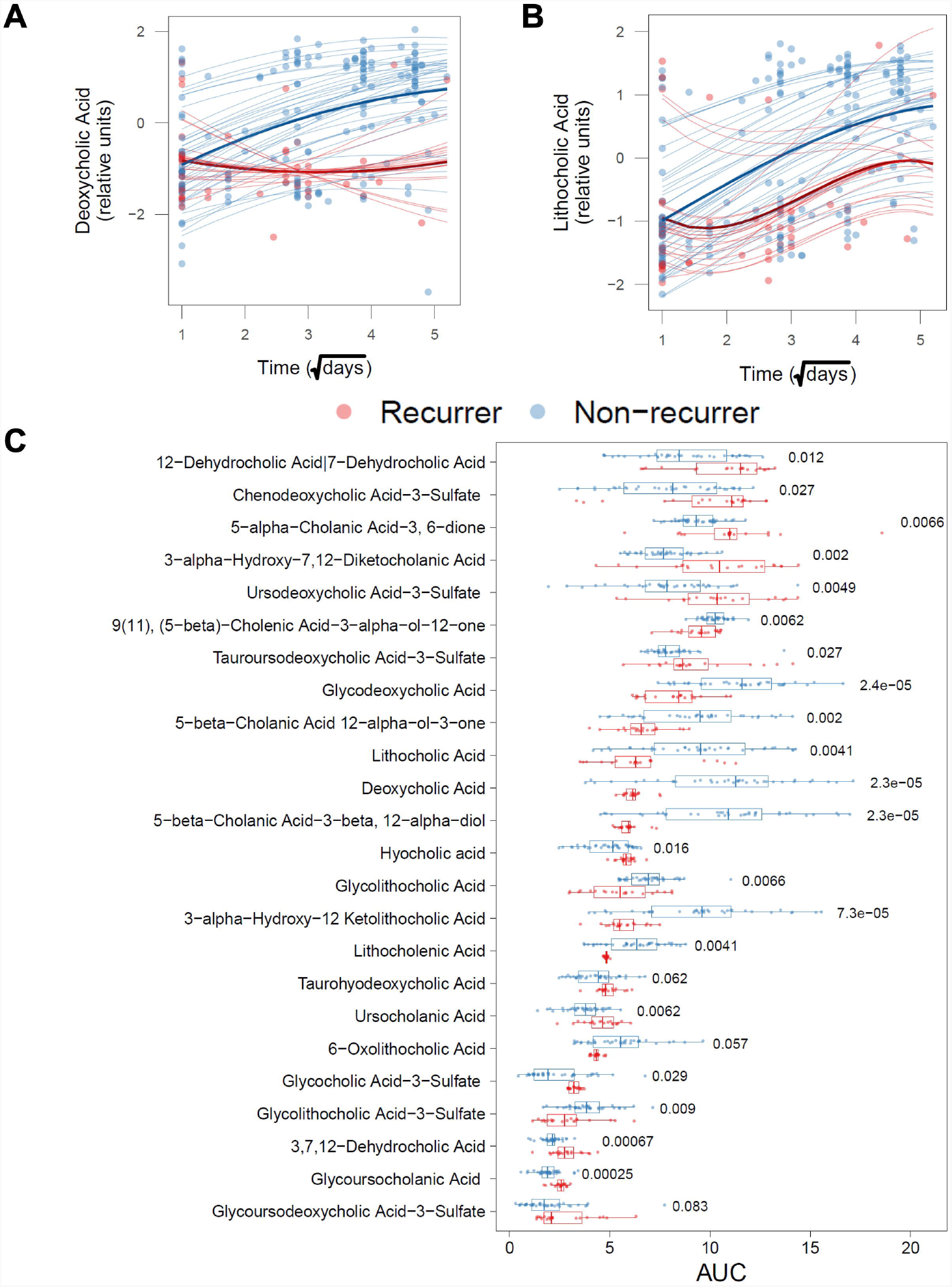
Longitudinal trajectories of fecal bile acids of primary CDI patients, comparing those experiencing recurrence versus those with no recurrence. As measured across days to weeks following the end of initial antibiotic therapy. Longitudinal trajectories are demonstrated for A) deoxycholic acid (DCA) and B) lithocholic acid (LCA); thick lines correspond to the mean fit for recurrers and non-recurrers, while thinner lines are the fitted individual trajectories, from which area under curve (AUC) was calculated. Dots correspond to the log_10_-transformed and mean-centered BA relative quantification. C) AUC of subject-specific trajectories ordered by increasing median in recurrers. *P* values were obtained using a Wilcoxon test and were adjusted (P*adj*) using the Benjamini-Hochberg method. Only significant features (P*adj* < 0.1) are shown.

Of the identified/ annotated bile acids, 24 were found to have marked differences in longitudinal trajectories, assessed via comparison of area under curve (AUC) of subject-specific trajectories among recurrence *vs* no recurrence (**Figure 2C** and **Supplementary Table 1**). Two of the fecal bile acids with the most marked difference in AUC between recurrers and non-recurrers were the secondary bile acids DCA and LCA, with AUCs for both significantly higher in the non-recurring patients (DCA: *P*_*adj*_= 2.3×10^−5^; LCA: *P*_*adj*_= 4.1×10^−3^, Mann-Whitney with Benjamini-Hochberg FDR; **Figure 2A**). Other fecal bile acid AUCs that strongly differentiated recurrence from non-recurrence (with larger AUC in non-recurrers) were particularly notable for also being secondary bile acids, and particularly derivatives of DCA and LCA. This included glycine-conjugated forms (glycoDCA and glycoLCA; *P*_*adj*_= 2.4×10^−5^ and 6.6×10^−3^ respectively, Mann-Whitney with Benjamini-Hochberg FDR), as well as forms presumably derived from microbial hydroxysteroid dehydrogenase (HSDH) activity: isoDCA (5-β-Cholanic Acid-3-β, 12-α-diol; *P*_*adj*_= 2.3×10^−5^), the 3β-HSDH-derived epimer of DCA; 3-oxoDCA (5-β-Cholanic Acid 12-α-ol-3-one; *P*_*adj*_*j*= 7.3×10^−5^), the 3α-HSDH-mediated dehydrogenation of DCA; and 12-ketoDCA (3-α-Hydroxy-12-ketolithocholic acid; *P*_*adj*_*j*= 7.3×10^−5^), the 12α-HSDH-mediated dehydrogenation of DCA. In contrast, participants experiencing recurrence (relative to those with non-recurrence) had higher AUC levels of primary bile acid derivatives, sulfated and/or amide-conjugated ursodeoxycholic acid (UDCA), or those similar to the 5β-cholanic acid (ursocholanic acid) skeleton from which all bile acids are derived; this included glycoursocholanic acid (*P*_*adj*_= 2.5×10^−5^), chenodeoxycholic acid-3-sulfate (*P*_*adj*_= 0.027), as well as a number of oxo-derivatives of cholic acid, i.e. 3,7,12-dehydrocholic acid, 3-α-hydroxy-7,12-diketocholanic acid, and 12-dehydrocholic acid|7-dehydrocholic acid (*P*_*adj*_= 6.7×10^−4^, 2×10^−3^ and 0.012 respectively).

Microbial bile salt hydrolases (BSH) are widely-distributed amongst bacteria resident in the gut, and their action (in removing the taurine or glycine groups of primary bile acids secreted into the gut) is the key rate-limiting step for microbiota-mediated 7α/β- dehydroxylation within the gastrointestinal tract [35]. Predicted bile salt hydrolase gene abundance has previously been demonstrated by our group to be reduced in stool in patients with recurrent CDI compared to those with primary CDI or control patients [15]; however, no comparison of BSH dynamics between recurring *vs*. non-recurring primary CDI patients has been described previously. As such, we undertook BSH activity assays on a subset of primary CDI patients who experienced CDI recurrence (*n*=9) and those with non-recurrence (*n*=18). BSH activity increased over time amongst patients who did not relapse (β= 0.056; likelihood ratio test *p*= 0.018; **Figure 3A**), while not enough data points were available to test changes in BSH activity over time in patients who relapsed. Comparison of BSH activity in recurrers *vs*. non-recurrers showed no significant difference in activity one day after antibiotic treatment cessation (Wilcoxon, *p*= 0.83), and a trend towards a lower BSH activity in patients who recurred at the latest timepoints measured, corresponding to a mean ± SD of 8 ± 7 days for recurrers and 35 ± 5 days for non-recurrers (Wilcoxon, *p*= 0.078; **Figure 3B**).

**Figure 3:**
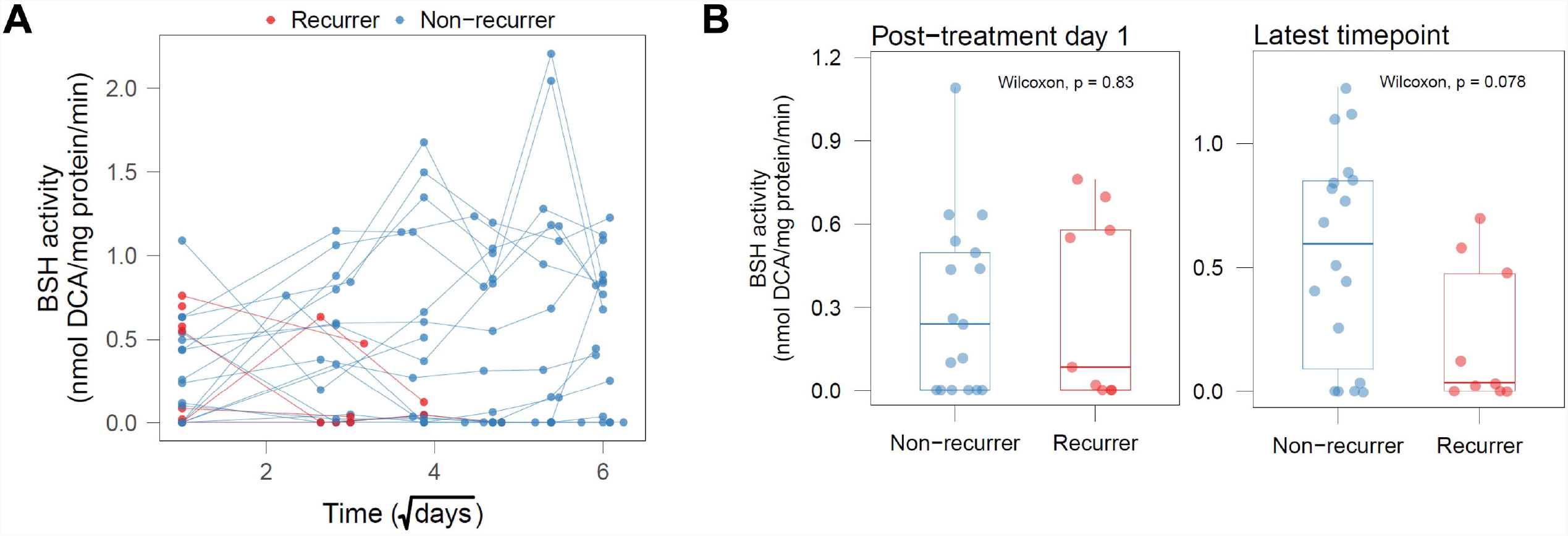
Comparative temporal dynamics of stool BSH activity changes in patients with primary CDI experiencing recurrence *versus* those with no recurrence. As assessed using plate-based precipitation assay. A) Time course of stool BSH activity. B) Comparison of stool BSH activity at one day post-treatment (left) and at the latest measured time point (right). P values were generated using Wilcoxon test (recurrers: *n*=9; non-recurrers: *n*=18).

### 4.4 Fecal bile acid profiles immediately following antibiotic treatment for primary CDI are related to risk of recurrence, and may be a predictive tool for recurrence

Comparison of AUC under the longitudinal trajectories allowed us to evaluate the overall recovery of bile acid species over a time interval, but it does not provide information on the imminent effect bile acids have on the risk of recurrence at a discrete timepoint. More specifically, it would be of clear clinical interest if it was feasible to use a ‘snapshot’ fecal bile acid profile obtained soon after the end of antibiotic treatment for primary CDI and use this to gain insight into future recurrence risk. As such, we sought to further investigate the relationship of specific bile acids to recurrence risk, and whether they had potential utility in use as a predictive marker of recurrence.

To explore the contribution of fecal bile acids to the risk of recurrence, we undertook a Bayesian joint longitudinal and survival modelling approach [34] (**Figure 4A** and **Supplementary Table 2**). We found 16 bile acids associated with increased risk of recurrence, while 15 were associated with reduced risk. Particularly noteworthy was the marked increased risk of recurrence associated with fecal ursocholanic acid (*p*=0.006), together with its conjugated derivatives, glycoursocholanic acid (*p*<0.001) and tauroursocholanic acid (*p*=0.034). In addition, UDCA and its sulfated and tauro-conjugated forms were also positively associated with recurrence. Conversely, it was noteworthy that those bile acids most strongly associated with a reduced risk of recurrence were predominantly glycine-conjugated primary bile acids, DCA and LCA, with many of them also sulfated (**Figure 4A**).

**Figure 4:**
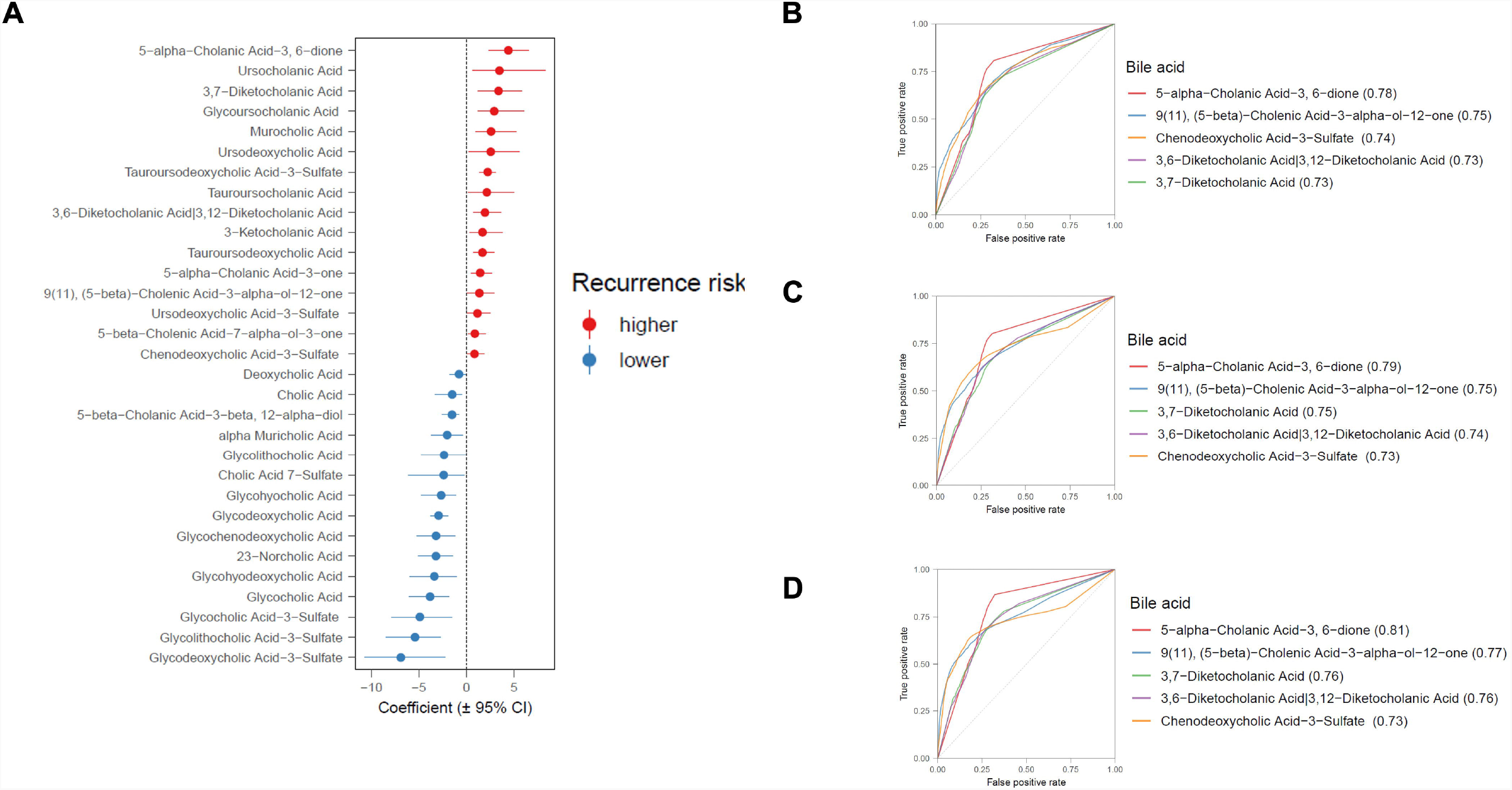
The contribution of fecal bile acids to risk of recurrence in patients with primary CDI, and their use as a predictive tool of recurrence. As assessed at early time points after completion of antibiotics as therapy for primary CDI. A) Bayesian joint longitudinal and survival model; a positive regression coefficient means that an increase in this particular bile acid will increase risk of recurrence, while a negative regression coefficient means that an increase in this bile acid will lower the risk of recurrence. CI denotes credibility interval. Longitudinal-survival model fitted to each bile acid was used to predict recurrence within 9 days post-CDI treatment using measurements at B) three timepoints; C) two time points; D) one time point only. Predictions were made using 80% of the data as training set and the other 20% as test set. Receiver operating characteristic plots show the mean of 1,000 Monte Carlo permutations for each bile acid, and the mean AUROC value is indicated in parenthesis in the figure legend.

We next aimed to assess the model’s performance in predicting recurrence in patients with primary CDI using fecal bile acid profiles. Given the time frame at which recurrences occur both within this study and in clinical practice more generally, we modelled prediction of recurrence within the 9 first days post-treatment, focusing on using only the measurements of post-treatment day 1, as it would be more translatable to clinical practice than conducting daily fecal samplings and BA measurements (**Figure 4D)**. Area under the receiver operating characteristic curve (AUROC) showed good prediction of recurrers within 9 days post-treatment by 5-α-cholanic acid-3, 6-dione (AUROC=0.81; **Figure 4D**). Four other fecal bile acids were also identified to have AUROCs > 0.73 for prediction of recurrence (**Figure 4D**). When using two or three consecutive measurements, 5-α-cholanic acid-3, 6-dione continued to be the best predictor of recurrence, but AUROC was slightly smaller (0.79 and 0.78, respectively) (**Figure 4B, 4C**), suggesting that measuring 5-α-cholanic acid-3, 6-dione on the first day post-treatment might be sufficient to predict the risk of recurrence.

## 5. Discussion

The body of evidence supporting a close interaction between perturbed gut microbiota-mediated bile acid metabolism and risk of CDI has been growing over the past decade [13], and particularly regarding the impact of enriched gut TCA and loss of gut secondary bile acids (particularly DCA and LCA) that characterise the CDI gut upon the ability of *C. difficile* to undergo germination, vegetative growth, and toxin activity. However, to date, relatively limited data has been described on the degree to which this axis is perturbed in primary CDI specifically, and – if so – whether it might be exploitable as a biomarker to predict recurrence within this condition.

Previous work from our laboratories described enrichment in gut primary bile acids and loss of secondary bile acids in patients with both primary and recurrent CDI, together with reduced predicted stool *bsh* gene abundance in recurrent CDI compared to primary CDI or controls; however, this was assessed only cross-sectionally rather than longitudinally [15]. One confounder in interpreting cross-sectional datasets in CDI (particularly primary CDI) is that vancomycin (the major anti-CDI therapy that most participants are initially treated with) has itself been associated with an altered gut microbiota profile, as well as by loss of fecal secondary bile acids and enrichment in primary unconjugated bile acids [36,37]. These data support and extend upon our previous results by demonstrating different dynamics of BSH functionality in recurrers compared to non-recurrers, as does the associated recovery of transition of primary to secondary bile acids and their derivatives (a further nuance in this system relates to the recent demonstration that different BSHs, due to their different substrate specificity, have different degrees of protective ability against CDI [38]). The conventional clinical interpretation of recurrence after an episode of primary CDI is to view this as a pathogen-derived event (e.g. related to re-exposure of a vulnerable host to *C. difficile*), and to respond with escalated or alternative anti-*C. difficile-*focused therapies; however, these data suggest that recurrence may alternatively be viewed as a failure of sufficient recovery of gut microbiome bile-metabolising functionality, and suggest by extension that targeted microbiome-focused therapies or bile acid co-treatments may have a potential role in mitigating the risk of recurrence. Supportive of this view, albeit from the context of recurrent rather than primary CDI, is the finding that a key mechanism underlying the efficacy of fecal microbiota transplant (FMT) in recurrent CDI is through restoration of microbial BSH functionality, and the associated restoration of a pre-morbid gut bile acid profile [18,39,40]. Similarly, both a spore-based ‘microbiome therapeutic’ derived from alcohol-shocked healthy donor stool and a live biotherapeutic product consisting of eight *Clostridia* strains were both shown to rapidly and sustainably restore stool secondary bile acids [41,42].

While much research to date on gut microbiome-bile acid interactions in the context of CDI has focused on the microbial enzymes BSH and 7-α-dehydroxylase [13], our data also give novel insight into disruption to complex additional interactions within this domain. For example, ursocholanic acid (also known as 5-β-cholanic acid, the ‘skeleton’ from which other bile acids are derived, lacking the characteristic bile acid hydroxyl groups at positions 3, 7 and 12) and its simple derivatives were demonstrated in both main models to be associated with adverse outcomes (i.e. higher AUCs in recurrers *vs* non-recurrers in the predicted individual longitudinal trajectories, and predictive of recurrence risk in the joint longitudinal-survival model). As another example, bile acids with oxo-/keto-groups appeared to associate with risk of recurrence across both models; this particularly applied to 3-oxo groups (presumably representing gut microbial 3α–hydroxysteroid dehydrogenase (HSDH) activity), with such bile acids more predictive of increased recurrence risk in the joint longitudinal-survival model. Conversely, one particularly interesting finding relates to isoDCA (5-β-cholanic acid-3-β, 12-α-diol), which was observed in both models to be associated with non-recurrence; this bile acid is the 3-β-hydroxy epimer of DCA, with its presence in stool likely resulting from metabolism of DCA first via bacterial 3α-HSDHs to 3-oxoDCA, before bacterial 3β-HSDH epimerization. As such, this suggests a role for gut microbial 3β-HSDH activity as being overall associated with protection from recurrence. This finding extends upon previous *in vitro* work demonstrating that a broad range of gut microbial derived secondary bile acids (including iso-forms) inhibit different aspects of the life cycle of *C. difficile* and its toxin activity [20]. The range of gut bacteria with 3α-HSDH functionality is focused around (but not based purely within) the class of *Clostridia*, and includes a number of bacteria with other well-characterised bile-metabolising functionality, e.g. *Clostridium scindens*, the archetypal bacterium recognised to possess 7-α-dehydroxylation functionality [43]; bacteria with 3β-HSDH functionality include *Ruminococcus gnavus* [44]. This observation leads us to conclude that the presence/ recovery of even very particular microbial bile-metabolising functions within the gut microbiota may be sufficient to profoundly affect clinical outcome. In particular, our data allows us to propose that primary CDI patients may typically retain gut bacterial 3-α-HSDH functionality, but only the additional presence of 3-β-HSDH functionality results in reduced recurrence risk; the presence of such functionality may represent gut microbiome recovery, and/or a gut environmental *milieu* supportive of activity of this enzyme (i.e. presence of required NADP(H), and appropriate pH and redox status [45]). The relevance of 3β-HSDH-mediated biotransformations of secondary bile acids to human health has been of recently-growing interest; one pertinent example of this is the recent demonstration that isoallolithocholic acid produced from this enzyme system by *Odoribacteriaceae* strains had potent effects against the growth of *C. difficile* and other pathobionts [46]. Furthermore, isoLCA has recently been described to suppress pro-inflammatory T_H_17 cell differentiation, and to be at reduced levels in patients with colitis (at least colitis caused by IBD) [47].

A further interesting observation relates to the presence of UDCA and its derivatives as associating with increased risk of recurrence in the joint longitudinal model; this seems initially counterintuitive, given the apparent benefits associated with the exogenous use of UDCA in a rodent CDI model (albeit potentially more through their impact upon bile acid receptor systems rather than via direct effects on the life cycle of *C. difficile* [48]). However, a possible explanation may be related to the observation from our longitudinal model that no bile acids with 7-α-hydroxyl groups had higher AUCs in patients experiencing recurrence as compared to non-recurrers. From these data, we conclude that recovery over time of the 7-α–dehydroxylase-driven transition from primary to secondary bile acids is a bacterial function that drives bile acid biotransformation in a way that reduces recurrence risk. By possessing a 7β group, UDCA is not readily acted upon by 7-α–dehydroxylase-producing bacteria, and may undergo downstream metabolism via more unconventional routes [49] such as isomerization back to 7α and then 7α–dehydroxylation, or direct 7β-dehydroxylation to LCA, that have less impact than immediate 7-α–dehydroxylation of primary bile acids upon direct bile acid-*C. difficile* interactions.

Extending upon this recognition of the centrality of microbiome-bile interactions in CDI outcomes - coupled with the current lack of clinical biomarkers to prognosticate recurrence in primary CDI – we further explored whether stool bile acid signatures may have utility as a predictive tool for recurrence of CDI. Work to date within this field is relatively limited, although we previously demonstrated that stool bile acid profiles could differentiate primary from recurrent CDI with 84.2% accuracy [15], and fecal LCA (combined with urinary p-cresol sulfate) has been demonstrated to predict response to FMT for recurrent CDI with 100% accuracy [50]. Our data here identifies at least five stool bile acids whose day 1 post-treatment measurement was enough to predict recurrence within 9 days with AUROC ranging from 0.81 to 0.73. This level of accuracy is similar to a previous study that used 16S rRNA gene sequencing of a single stool sample from each of 88 primary CDI patients to derive a microbial signature ‘risk index’ to predict recurrence, where an AUROC of 0.78 for prediction of recurrence was reported [51]. Collectively, data such as these suggest that microbiome and metabolome profiling in primary CDI may hold promise as accurate biologically-rationale biomarkers to predict recurrence. However, such studies would need replication in larger cohorts before they could be considered for use within clinical practice, and pragmatic aspects regarding use of such assays (i.e. time consuming multi-step aspect of sample preparation and batch effects in mass spectrometry analysis) would also require careful consideration. Regarding the actual bile acids that predicted recurrence, it was of particular interest that an unusual ‘allo’ bile acid - 5-α-cholanic acid-3, 6-dione - was the most robust predictor of recurrence, especially since formation of allo bile acids is recognised to be a function of the gut microbiome of particular mammals (including rabbits and rates) but not of humans [52]; further work is needed to establish the potential biological significance of this bile acid to this disease state.

In conclusion, our data demonstrate distinctive differential gut bile acid trajectories that differentiate recurrence from non-recurrence in patients with primary CDI, and define the degree to which different gut bile acids associate with recurrence risk. These data extend upon existing knowledge regarding the influence of microbiome-mediated bile acid metabolism pathways upon clinical outcomes in CDI. By extension, these data suggest a clinical case for focus upon therapies targeted at recovery of microbiome-bile acid interactions to potentially reduce the risk of recurrence after primary CDI, and a feasible role for stool bile acids as novel, rationale biomarkers to predict CDI recurrence.

## Data Availability

The bile acid profiling data generated for this project is available on the MetaboLights Repository, study number: MTBLS657. All data produced in the present study are available upon reasonable request to the authors.

https://www.ebi.ac.uk/metabolights/

## Acknowledgements

Nil.

