## Supplementary Material for "Fecal bile acid profiles predict recurrence in patients with primary *Clostridioides difficile* infection"

**Supplementary Tables:**

**Supplementary Table 1: Full results from longitudinal trajectory analysis.** AUC of predicted within-individual trajectories were compared between recurrers and non-recurrers. *p* values were obtained using a Mann-Whitney test and were adjusted using the Benjamini-Hochberg method. Mass: charge ratio (*m/z*) and retention time of these bile acids as previously-described [1]. Bile acids sorted by *Padj.*

| **Name of bile acid** | **Mean (non-recurrer)** | **Mean (recurrer)** | ***P*** | ***Padj*** |
| --- | --- | --- | --- | --- |
| 5-beta-Cholanic Acid-3-beta, 12-alpha-diol (isodeoxycholic acid) | 10.72502017 | 5.922005944 | 3.58E-07 | 2.30E-05 |
| Deoxycholic Acid | 10.74470093 | 6.188443722 | 6.47E-07 | 2.30E-05 |
| Glycodeoxycholic Acid | 11.42855772 | 8.193628909 | 1.02E-06 | 2.40E-05 |
| 3-alpha-Hydroxy-12 Ketolithocholic Acid | 9.477119413 | 5.671141516 | 4.15E-06 | 7.30E-05 |
| Glycoursocholanic Acid | 1.94112087 | 2.505561591 | 1.80E-05 | 0.00025 |
| 3,7,12-Dehydrocholic Acid | 2.096531707 | 2.834345148 | 5.72E-05 | 0.00067 |
| 3-alpha-Hydroxy-7,12-Diketocholanic Acid | 7.752245659 | 10.31615358 | 0.000225716 | 0.002 |
| 5-beta-Cholanic Acid 12-alpha-ol-3-one | 9.261802435 | 6.615799842 | 0.000225716 | 0.002 |
| Lithocholenic Acid | 6.254435738 | 4.825502669 | 0.000586087 | 0.0041 |
| Lithocholic Acid | 9.536265973 | 6.663775393 | 0.000586087 | 0.0041 |
| Ursodeoxycholic Acid-3-Sulfate | 7.947897868 | 10.38786479 | 0.000774041 | 0.0049 |
| 9(11), (5-beta)-Cholenic Acid-3-alpha-ol-12-one | 10.25832278 | 9.439844543 | 0.001159659 | 0.0062 |
| Ursocholanic Acid | 3.681600935 | 4.552109889 | 0.001159659 | 0.0062 |
| 5-alpha-Cholanic Acid-3, 6-dione | 9.355459137 | 10.96817292 | 0.001411375 | 0.0066 |
| Glycolithocholic Acid | 6.986789736 | 5.492292652 | 0.001411375 | 0.0066 |
| Glycolithocholic Acid-3-Sulfate | 3.928233478 | 2.915522215 | 0.002067588 | 0.009 |
| 12-Dehydrocholic Acid\|7-Dehydrocholic Acid | 8.667779026 | 10.68146397 | 0.002811148 | 0.012 |
| Hyocholic acid | 4.905341305 | 5.834579324 | 0.004012738 | 0.016 |
| Chenodeoxycholic Acid-3-Sulfate | 7.953296006 | 9.822540041 | 0.007854206 | 0.027 |
| Tauroursodeoxycholic Acid-3-Sulfate | 8.074794201 | 9.282215581 | 0.007441514 | 0.027 |
| Glycocholic Acid-3-Sulfate | 2.428568495 | 3.216663139 | 0.008740241 | 0.029 |
| 6-Oxolithocholic Acid | 5.466052868 | 4.333058576 | 0.017770839 | 0.057 |
| Taurohyodeoxycholic Acid | 4.308697344 | 4.904378335 | 0.020513406 | 0.062 |
| Glycoursodeoxycholic Acid-3-Sulfate | 1.973619823 | 2.722796326 | 0.028350232 | 0.083 |
| Glycohyocholic Acid | 4.648709224 | 5.369723284 | 0.065710074 | 0.18 |
| 5-alpha-Cholanic Acid-3-one | 9.30146241 | 8.587522222 | 0.088970662 | 0.24 |
| Taurodeoxycholic Acid-3-Sulfate | 2.844379347 | 3.129814026 | 0.106536508 | 0.28 |
| Glycochenodeoxycholic Acid | 4.5679842 | 3.989314058 | 0.140106038 | 0.35 |
| Cholic Acid 7-Sulfate | 2.203909859 | 2.051370817 | 0.192599271 | 0.45 |
| Glycocholic Acid | 6.142129971 | 5.549564056 | 0.192599271 | 0.45 |
| Taurocholic Acid-3-Sulfate | 6.909485039 | 6.882820412 | 0.204610971 | 0.46 |
| Isolithocholic Acid | 10.89188353 | 9.989321423 | 0.217148114 | 0.48 |
| 3-Dehydrocholic Acid | 5.395383631 | 6.120025543 | 0.236954957 | 0.5 |
| Glycodeoxycholic Acid-3-Sulfate | 3.291403717 | 3.260862708 | 0.265264156 | 0.55 |
| 23-Norcholic Acid | 3.254433043 | 2.900531667 | 0.337038355 | 0.6 |
| 5-beta-Cholanic Acid-3-alpha, 6-alpha-diol-7-one | 12.90805388 | 12.6733486 | 0.311864393 | 0.6 |
| Cholic Acid-3-Sulfate | 14.04348085 | 14.52863652 | 0.328508271 | 0.6 |
| Glycohyodeoxycholic Acid | 3.684965104 | 3.894243555 | 0.311864393 | 0.6 |
| Taurochenodeoxycholic Acid-3-Sulfate | 5.313786576 | 5.821196781 | 0.337038355 | 0.6 |
| Tauroursocholanic Acid | 1.948690521 | 2.072642765 | 0.345706919 | 0.6 |
| 3-alpha-Hydroxy-7 Ketolithocholic Acid | 7.141532255 | 7.557585533 | 0.363458134 | 0.61 |
| 5-beta-Cholenic Acid-7-alpha-ol-3-one | 6.900167155 | 7.471914173 | 0.354513644 | 0.61 |
| 3,7,12-Taurodehydrocholic Acid | 2.622204861 | 2.491599241 | 0.429875579 | 0.65 |
| 5-alpha-Cholanic Acid-3-alpha-ol-6-one | 5.072385557 | 4.768598158 | 0.41998507 | 0.65 |
| Chenodeoxycholic Acid | 13.68272216 | 12.87091212 | 0.429875579 | 0.65 |
| Taurocholic Acid | 5.736322214 | 6.522678581 | 0.400602786 | 0.65 |
| Glycoursodeoxycholic Acid | 5.823923408 | 6.468388901 | 0.439897577 | 0.66 |
| Tauro omega-Muricholic Acid | 6.174683637 | 6.509927768 | 0.450049887 | 0.66 |
| Isoallolithocholic Acid | 7.101190994 | 6.509101222 | 0.502718756 | 0.7 |
| omega Muricholic Acid | 6.262658923 | 6.15665648 | 0.502718756 | 0.7 |
| Taurolithocholic Acid | 3.344328114 | 2.92632461 | 0.547046992 | 0.75 |
| alpha Muricholic Acid | 5.472489607 | 5.646936157 | 0.641013424 | 0.83 |
| Deoxycholic Acid-3-Sulfate | 6.246576474 | 5.990823851 | 0.653211204 | 0.83 |
| Lithocholic Acid 3-Sulfate | 13.71089313 | 13.2937248 | 0.628910092 | 0.83 |
| Taurohyocholic Acid | 2.694577381 | 3.036011785 | 0.641013424 | 0.83 |
| Glycochenodeoxycholic Acid-3-Sulfate | 3.003211627 | 3.191173986 | 0.728222223 | 0.91 |
| 3-Ketocholanic Acid | 9.36686536 | 9.687277553 | 0.766764463 | 0.93 |
| 3,6-Diketocholanic Acid\|3,12-Diketocholanic Acid | 12.65406097 | 12.83561321 | 0.766764463 | 0.93 |
| 3,7-Diketocholanic Acid | 9.894219497 | 10.35669898 | 0.792784488 | 0.94 |
| 5-Cholenic Acid-3-beta-ol | 8.171453929 | 8.187680489 | 0.845482409 | 0.98 |
| beta Muricholic Acid | 5.78031547 | 5.837045116 | 0.925746246 | 0.98 |
| Cholic Acid | 21.62917389 | 20.81050636 | 0.925746246 | 0.98 |
| Murocholic Acid | 6.296357802 | 5.991441925 | 0.966210231 | 0.98 |
| Tauro-beta Muricholic Acid \|Tauro-alpha Muricholic Acid | 3.87942197 | 3.784509436 | 0.979722328 | 0.98 |
| Taurochenodeoxycholic Acid | 4.876523426 | 5.024678888 | 0.979722328 | 0.98 |
| Taurodeoxycholic Acid | 4.028487587 | 3.730629428 | 0.939218398 | 0.98 |
| Taurolithocholic Acid 3-Sulfate | 3.024639479 | 2.992934478 | 0.979722328 | 0.98 |
| Tauroursodeoxycholic Acid | 5.340776034 | 5.835282131 | 0.91229497 | 0.98 |
| Ursocholic Acid | 21.62753042 | 20.808975 | 0.925746246 | 0.98 |
| Ursodeoxycholic Acid | 8.765310364 | 8.663687596 | 0.858773422 | 0.98 |

**Supplementary Table 2: Full results from joint longitudinal and survival modelling approach.** Bayesian joint longitudinal-survival model was fitted to each bile acid. Table shows the summary of the obtained survival regression coefficients (θ), with the 95% credibility interval (CI) and the P value, calculated as 2xmin{P(θ > 0), P(θ < 0)}. Mass: charge ratio (*m/z*) and retention time of these bile acids as previously-described [1]. Bile acids sorted by coefficient value.

| name | Value | 2.5% CI | 97.5% CI | P | Signif |
| --- | --- | --- | --- | --- | --- |
| Glycodeoxycholic Acid-3-Sulfate | -6.894473352 | -10.7084 | -2.24696 | 0.002 | * |
| Glycolithocholic Acid-3-Sulfate | -5.396359367 | -8.47235 | -2.70466 | <0.001 | * |
| Glycocholic Acid-3-Sulfate | -4.89231351 | -7.85217 | -1.53056 | <0.001 | * |
| Glycocholic Acid | -3.815195326 | -5.98806 | -1.82793 | <0.001 | * |
| Glycohyodeoxycholic Acid | -3.367089464 | -5.95279 | -1.03873 | 0.001 | * |
| 23-Norcholic Acid | -3.200859131 | -5.02063 | -1.42789 | <0.001 | * |
| Glycochenodeoxycholic Acid | -3.191789273 | -5.23458 | -1.16992 | 0.001 | * |
| Glycodeoxycholic Acid | -2.922399507 | -3.77866 | -1.89924 | <0.001 | * |
| Glycohyocholic Acid | -2.6479685 | -4.74822 | -1.09652 | <0.001 | * |
| Cholic Acid 7-Sulfate | -2.385904584 | -6.07499 | -0.21513 | 0.034 | * |
| Glycolithocholic Acid | -2.350322701 | -4.71233 | -0.02013 | 0.048 | * |
| alpha Muricholic Acid | -2.017779173 | -3.72678 | -0.40554 | 0.011 | * |
| 5-beta-Cholanic Acid-3-beta, 12-alpha-diol | -1.50669788 | -2.52703 | -0.75167 | <0.001 | * |
| Cholic Acid | -1.490611846 | -3.30394 | -0.43986 | 0.005 | * |
| Deoxycholic Acid | -0.772198607 | -1.74563 | -0.07265 | 0.033 | * |
| Chenodeoxycholic Acid-3-Sulfate | 0.858898912 | 0.129307 | 1.849439 | 0.019 | * |
| 5-beta-Cholenic Acid-7-alpha-ol-3-one | 0.895448048 | 0.165871 | 1.99734 | 0.02 | * |
| Ursodeoxycholic Acid-3-Sulfate | 1.166440649 | 0.137408 | 2.48949 | 0.026 | * |
| 9(11), (5-beta)-Cholenic Acid-3-alpha-ol-12-one | 1.37350217 | 0.074742 | 2.912029 | 0.038 | * |
| 5-alpha-Cholanic Acid-3-one | 1.457853155 | 0.482328 | 2.645038 | 0.002 | * |
| Tauroursodeoxycholic Acid | 1.697813163 | 0.781243 | 2.941096 | <0.001 | * |
| 3-Ketocholanic Acid | 1.702629448 | 0.358398 | 3.827164 | 0.006 | * |
| 3,6-Diketocholanic Acid\|3,12-Diketocholanic Acid | 1.958212102 | 0.721592 | 3.631719 | <0.001 | * |
| Tauroursocholanic Acid | 2.157726073 | 0.150233 | 5.023054 | 0.036 | * |
| Tauroursodeoxycholic Acid-3-Sulfate | 2.249795519 | 1.407537 | 3.05608 | <0.001 | * |
| Ursodeoxycholic Acid | 2.56934301 | 0.286281 | 5.558151 | 0.029 | * |
| Murocholic Acid | 2.601461824 | 0.963957 | 5.276511 | 0.004 | * |
| Glycoursocholanic Acid | 2.934481256 | 1.24894 | 6.041247 | <0.001 | * |
| 3,7-Diketocholanic Acid | 3.387390958 | 1.226572 | 5.787827 | <0.001 | * |
| Ursocholanic Acid | 3.483933979 | 0.660176 | 8.345488 | 0.006 | * |
| 5-alpha-Cholanic Acid-3, 6-dione | 4.42469188 | 2.337682 | 6.541606 | <0.001 | * |
| Taurodeoxycholic Acid-3-Sulfate | -2.563756568 | -7.60292 | 0.815676 | 0.145 |  |
| Lithocholic Acid 3-Sulfate | -1.895931515 | -4.00878 | 0.339642 | 0.082 |  |
| Glycochenodeoxycholic Acid-3-Sulfate | -1.375791626 | -5.83116 | 0.234162 | 0.202 |  |
| 5-alpha-Cholanic Acid-3-alpha-ol-6-one | -1.012229737 | -2.63441 | 0.369581 | 0.163 |  |
| Lithocholic Acid | -0.850218467 | -3.28627 | 0.282579 | 0.158 |  |
| Isolithocholic Acid | -0.592485928 | -1.34661 | 0.178985 | 0.118 |  |
| Taurocholic Acid-3-Sulfate | -0.449348081 | -1.91773 | 1.082158 | 0.511 |  |
| 3-alpha-Hydroxy-12 Ketolithocholic Acid | -0.401201356 | -1.30564 | 0.311838 | 0.299 |  |
| Ursocholic Acid | -0.377434328 | -1.25696 | 0.608781 | 0.407 |  |
| Taurochenodeoxycholic Acid-3-Sulfate | -0.284307553 | -0.84288 | 0.296302 | 0.282 |  |
| Taurodeoxycholic Acid | -0.275720285 | -1.0712 | 0.43799 | 0.469 |  |
| Taurohyocholic Acid | -0.267890833 | -1.01628 | 0.523948 | 0.44 |  |
| Chenodeoxycholic Acid | -0.213725244 | -1.34481 | 0.756082 | 0.714 |  |
| Lithocholenic Acid | -0.209702915 | -1.47696 | 1.107022 | 0.724 |  |
| 3-alpha-Hydroxy-7,12-Diketocholanic Acid | -0.183371674 | -0.78238 | 0.455592 | 0.513 |  |
| Taurochenodeoxycholic Acid | -0.167674174 | -0.8356 | 0.456687 | 0.575 |  |
| 5-beta-Cholanic Acid 12-alpha-ol-3-one | -0.10829982 | -1.61597 | 0.755993 | 0.964 |  |
| 5-Cholenic Acid-3-beta-ol | -0.09247125 | -0.81037 | 0.638028 | 0.801 |  |
| Taurocholic Acid | -0.084574904 | -0.63428 | 0.495012 | 0.744 |  |
| Tauro-beta Muricholic Acid \|Tauro-alpha Muricholic Acid | -0.024384315 | -0.93273 | 0.975541 | 0.948 |  |
| Tauro omega-Muricholic Acid | 0.036367106 | -0.55016 | 0.733753 | 0.974 |  |
| Taurolithocholic Acid | 0.102239311 | -1.10921 | 2.081165 | 0.989 |  |
| Deoxycholic Acid-3-Sulfate | 0.20070966 | -0.80106 | 2.234009 | 0.779 |  |
| 3,7,12-Taurodehydrocholic Acid | 0.334612821 | -4.00765 | 2.117522 | 0.49 |  |
| 5-beta-Cholanic Acid-3-alpha, 6-alpha-diol-7-one | 0.365639123 | -0.82065 | 1.948148 | 0.631 |  |
| omega Muricholic Acid | 0.508528753 | -0.9745 | 1.958404 | 0.511 |  |
| Glycoursodeoxycholic Acid-3-Sulfate | 0.671971031 | -0.4553 | 1.818971 | 0.194 |  |
| 3,7,12-Dehydrocholic Acid | 0.723378177 | -0.3395 | 1.837567 | 0.142 |  |
| 6-Oxolithocholic Acid | 0.741713265 | -0.53853 | 2.629264 | 0.305 |  |
| Isoallolithocholic Acid | 0.917168397 | -0.19709 | 2.048069 | 0.093 |  |
| 3-Dehydrocholic Acid | 0.923526028 | -0.90214 | 2.815678 | 0.309 |  |
| Hyocholic acid | 1.090735226 | -0.23751 | 4.770866 | 0.124 |  |
| beta Muricholic Acid | 1.091531864 | -0.14986 | 2.247725 | 0.081 |  |
| Glycoursodeoxycholic Acid | 1.253499084 | -0.43107 | 3.996896 | 0.095 |  |
| Cholic Acid-3-Sulfate | 1.335545123 | -0.46307 | 3.601821 | 0.168 |  |
| Taurolithocholic Acid 3-Sulfate | 1.557223723 | -10.1581 | 10.54434 | 0.748 |  |
| 12-Dehydrocholic Acid\|7-Dehydrocholic Acid | 1.657503384 | -0.2656 | 4.0532 | 0.164 |  |
| Taurohyodeoxycholic Acid | 1.713472889 | -0.93 | 4.096511 | 0.218 |  |
| 3-alpha-Hydroxy-7 Ketolithocholic Acid | 1.825529191 | -0.48899 | 5.584918 | 0.527 |  |

**Supplementary Figures:**

**Supplementary Figure 1. Dynamics of recurrence.** A) Recurrence event for each patient who experienced a recurrence (*n*=20) with time as days (top) or as the square root of days (bottom). Red (top) or green (bottom) dots and labels indicate the time of recurrence while black dots indicate the times when a fecal sample was obtained. B) Deoxycholic acid (left) and lithocholic acid (right) individual trajectories across time in days (top) or in squared root of days (bottom). The y-axis shows log10-transformed and mean-centered relative intensities.

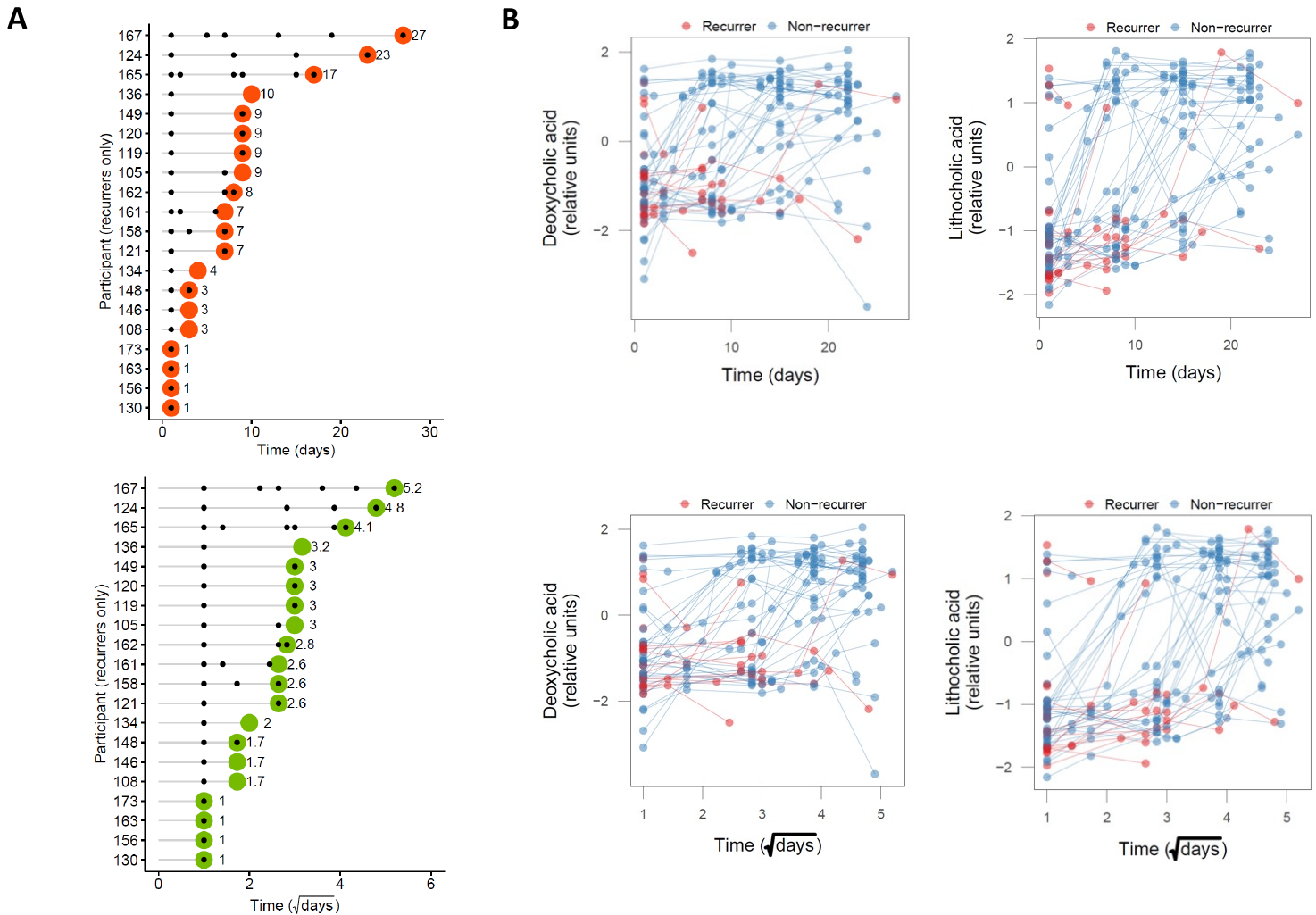
